# Developing an Indicator for Age-Friendly Communities: The Japan Gerontological Evaluation Study

**DOI:** 10.1101/2024.05.17.24307523

**Authors:** Satoko Fujihara, Taiji Noguchi, Kazushige Ide, Seungwon Jeong, Katsunori Kondo, Toshiyuki Ojima

## Abstract

**Background:** Age-friendly communities (AFCs) aim to create inclusive societies for older people. Despite the World Health Organization (WHO)’s emphasis on incorporating dementia-friendliness across all phases, including planning, implementation, monitoring, evaluation, and scale-up, there are very few community-level indicators that incorporate dementia-friendly elements.

**Objective:** To develop a community-level AFC indicator based on WHO AFC guidelines incorporating dementia-friendly elements, and examine its validity and reliability.

**Design:** A repeated cross-sectional study using data from the 2016 and 2019 waves of the Japan Gerontological Evaluation Study.

**Setting and Subjects:** Data were collected from 61 school districts in 16 Japanese municipalities, involving 45,162 individuals aged ≥65 years in 2016, and 39,313 in 2019. The 2016 and 2019 datasets were the development and retest samples, respectively.

**Methods:** After identifying 23 candidate items according to the WHO AFC guidelines and expert reviews, data were aggregated by school district. Exploratory factor analysis on the 2016 data helped derive factor structure, confirming reproducibility with the 2019 data. Internal consistency and test-retest reliability were evaluated.

**Results:** The final 17-item indicator comprised three subscales: *Social inclusion and dementia-friendliness* (7 items, α = 0.86), *Social engagement and communication* (5 items, α = 0.78), and *Age-friendly physical environment* (5 items, α = 0.82). The structure showed adequate test-retest reliability (r = 0.71–0.79; ICC = 0.67–0.78).

**Conclusions:** A valid and reliable 17-item community-level indicator was developed, which aligns with the WHO framework and also incorporates dementia-friendly elements. This indicator is useful for monitoring and evaluating to promote the AFC and dementia-friendly communities.

## 1. Introduction

The number of people aged 65 years or older globally was approximately 761 million in 2021 and is predicted to more than double by 2050 to 1.6 billion [1]. Simultaneously, urbanization is accelerating, with 55% of the global population residing in urban areas as of 2018; this number is projected to grow to 68% by 2050. Rapid urbanization presents risks to health, society, and the environment [2, 3], often adversely affecting the health and well-being of older people and limiting their ability to meet their basic needs, build and maintain relationships, and take decisions. Therefore, creating age-friendly communities (AFCs) in cities is crucial to ensure quality of life and dignity for older people. The World Health Organization (WHO) defines an AFC as a community that promotes active, healthy aging, and has provided guidelines to help cities plan for rapid population aging [4].

In 2020, the WHO and United Nations Member States launched the UN Decade of Healthy Aging to help people live longer, irrespective of their location [5]. In particular, the development of healthy aging environments, such as AFCs, is attracting attention as an important social issue. The AFC framework proposed by the WHO comprises eight domains: outdoor spaces and buildings, transportation, housing, social participation, respect and social inclusion, civic participation and employment, communication and information, and community support and health services [4]. To put this framework into practice, a robust monitoring and evaluation system is essential [3]. Monitoring refers to the ongoing collection of data to determine the progress of activities, such as improvements in public transportation for older people and recreation programs. Evaluation refers to assessing whether desired outcomes have been achieved, such as whether public transport improvements or recreational programs have positively impacted the physical health of older people [3]. For effective monitoring and evaluation of AFCs, a robust age-friendly indicator is indispensable.

The development of indicators evaluating AFCs has mainly been based on the WHO AFC guidelines [6-10]. Recent studies have developed individual-level AFC indicators in countries such as the United Kingdom [9], the Netherlands [10], the United States [11], and Turkey [12], and tested their validity and reliability. For example, in Turkey [12], an indicator with 20 items spanning eight domains was developed and checked for validity and reliability among 306 older people, following Dikkenn et al. [10]. Meanwhile, a few community-level AFC indicator was developed [13, 14]. Based on the Prospective Urban Rural Epidemiology (PURE) study [15], which includes participants in low-, middle-, and high-income countries, Rugel Raham et al. [14] surveyed people aged 35–70 years in urban and rural communities in 20 countries and developed community-level healthy-aging indicator.

AFCs cannot be isolated from personal factors such as age, gender, income, and functional status, as well as other levels of influence such as the environment [16]. Most of the above-mentioned indicators are at the individual-level [9-12], meaning that only a limited number of community-level indicators [13, 14]. The monitoring and evaluation systems for AFC programmes are not aligned with local, regional, national, or global monitoring and evaluation frameworks, resulting in poor coordination [3]. Hence, it is important to develop indicators suitable for each country or area that can be shared across sectors and compared across regions.

With the global population aging and over 55 million individuals living with dementia worldwide, the concept of dementia-friendly communities (DFCs) has emerged to support people with dementia and their caregivers [17]. DFCs and AFCs share some common goals, such as enabling older people to maintain independence and fostering a supportive environment through stakeholder engagement [18-20]. However, though similar, they are not the same in every respect [19, 20]. AFCs take a whole-person view of older individuals, whereas dementia-friendly initiatives focus specifically on dementia, addressing its unique challenges [20]. As Turner [21] points out, rather than competing, AFCs and DFCs can expand their reach and impact as complementary initiatives, benefiting millions. The WHO guidelines (2021) [18] emphasize the importance of dementia-friendly initiatives working in harmony with AFC-related initiatives. According to these guidelines, an AFC must comprehensively integrate dementia-friendliness across all phases, including planning, implementation, monitoring, evaluation, and scale-up [18]. Furthermore, the WHO underscores the necessity of adapting dementia-friendly initiatives within the broader AFC context when required, specifically meeting the unique needs of those with dementia and their families or caregivers [18]. However, there are few community-level AFC indicators that incorporate dementia-friendly elements.

The objectives of this study were twofold: 1) to develop a community-level AFC indicator grounded in the WHO AFC guidelines incorporating considerations of dementia-friendly elements, and 2) to examine the validity, and reliability of the developed indicator to assess temporal stability.

## 2. Methods

### 2.1 Data

We used repeated cross-sectional data derived from the 2016 and 2019 waves of the Japan Gerontological Evaluation Study (JAGES). The JAGES is an ongoing cohort study investigating social and behavioural factors related to health decline, including mortality and the onset of functional or cognitive impairment among individuals aged 65 years or older [22]. We used the 2016 wave as the development sample and the 2019 wave as the retest sample.

The development sample comprised self-administered questionnaires mailed in October and November 2016 to independent older people aged 65 years and over, who were not eligible for long-term care insurance benefits. The questionnaires were mailed to 115,350 people living in 250 communities, defined by school district, in 16 municipalities in 8 prefectures. The questionnaires were collected from 81,515 people (response rate: 70.7%). The valid responses respondents were 65,722 individuals. The survey consisted of two parts: a common core item administered to all respondents and eight randomly distributed modules. The development sample used data from responses to the core items and three module items from the eight models, including the AFC and DFC indicators. The school district was adopted as the community unit, and to avoid inaccuracies due to sample size, school districts with fewer than 30 respondents were excluded, resulting in 62 school districts with 45,503 respondents. The selection of school districts as the foundational community unit was predicated on their geographical suitability for older people, who are able to easily move around in these areas by foot or bicycle. Additionally, the presence of numerous local activities in these districts, such as senior citizen clubs and sports organizations, underscores their significance as integral components for the evaluation of local public health initiatives [23].

The retest sample similarly included a self-administered postal survey of independent older people aged 65 and over, who were not eligible for long-term care insurance benefits, between November 2019 and January 2020. The retest sample used data from responses to the core items and one module item from each of the eight models, including the AFC and DFC indicators. As in the development sample, we excluded areas with school districts with fewer than 30 respondents, resulting in 68 school districts with 40,998 respondents.

Note that we included school districts in these samples by excluding less than 30 school districts that were not covered by each other. Therefore, finally, we excluded school districts in the development sample that were not covered by the retest sample and included those in the retest sample that were not covered by the development sample.

Thus, the final analysis included 61 school districts and 45,162 respondents in 2016 and 39,313 respondents in 2019 (Figure 1). The JAGES protocol was approved by the Ethical Committee of Chiba University (approval no. M10460). The self-administered questionnaire was accompanied by a description of the study, and the return of the completed questionnaire was regarded as the participant’s provision of informed consent.

**Figure. 1.**
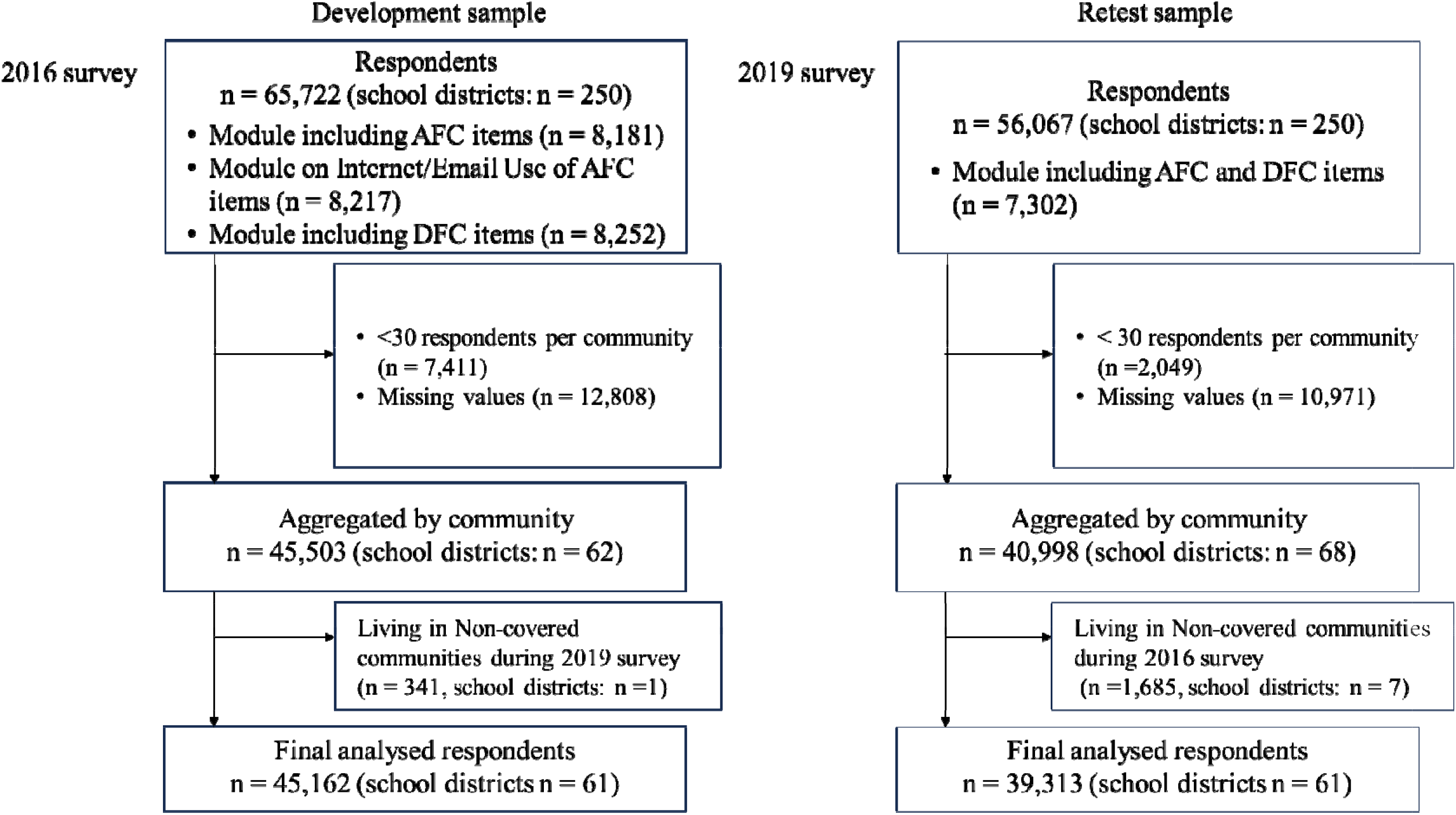
Flowchart of the study population: JAGES 2016 survey and 2019 survey.

[Figure 1 approximately here]

### 2.2 Selection of Candidate Indicators for Age-Friendly Communities

This study created a set of 23 candidate items reflecting the concept of age-friendly communities based on the eight indicators defined in the WHO guide entitled ‘Measuring the Age-friendliness of Cities’ [24]. Furthermore, based on previous dementia-related research [25-27], dementia-friendly elements related to the social environment were added. To assess the indicators’ content validity, several gerontology experts (a geriatrician, gerontological nurse, geriatric physiotherapist, healthcare official, and welfare management official) were consulted, who helped refine the list to 23 items. During a monthly research meeting hosted by the JAGES office, the experts reviewed the indicators and concluded that they effectively assess age- and dementia-friendliness in the social environment context. The 23 items spanned nine domains including the eight WHO domains and DFC. eTable1 shows detailed information on the 23-items. Higher composite scores indicate greater age-friendliness aggregated at the community level.

### 2.3 Statistical Analysis

First, for both the development and retest samples, the mean and standard deviation were computed for each item. Subsequently, an exploratory factor analysis (EFA) was conducted on the development samples to evaluate construct validity. This process began with an examination of the inter-item correlations. The appropriate number of factors to retain was determined using multiple criteria: Kaiser’s criterion of eigenvalues exceeding unity, inspection of the scree plot, and parallel analysis. The EFA employed the maximum likelihood estimation method, coupled with promax rotations. Factor loadings below the 0.4 threshold were eliminated. To assess the replicability of the factor structure identified within the development samples, EFA was subsequently performed on the retest samples. For both the development and retest samples, the internal consistency reliability of the indicators was evaluated by calculating Cronbach’s alpha coefficients. Additionally, subscale scores were calculated for the two samples by averaging each item in each subscale of the extracted factor structure. To examine the stability of the developed scale over time, test returns were evaluated using Pearson correlation coefficients for each subscale score in the two time series samples and the intraclass correlation coefficient (ICC), calculated as ICC [2,1]. All statistical analyses were conducted using R software (Version 4.3.0 for Windows; R Foundation for Statistical Computing, Vienna, Austria).

## 3. Results

Table 1 shows the characteristics of the candidate items for the development samples in 2016 and retest samples in 2019. ‘Participation in learning or cultural group’ had the same proportion between the development and retest samples. By contrast, ‘use of the Internet or email’ increased by 13.0% and ‘understanding of people with dementia’ decreased by 8.7%. eTable 2 shows the correlation coefficients of the development samples in 2016. Table 2 shows the results of EFA. It details the exclusion of the six items—housing type, participation in senior citizen club, participation in paid work, awareness of living with dementia, understanding of people with dementia, and decision-making support for people with dementia—owing to low factor loadings (less than 0.40). Therefore, 17 items were finally adopted, with a three-factor structure. Factor 1 – *Social inclusion and dementia-friendliness* comprises seven items and is strongly associated with items such as ‘sense of belonging to the community’, ‘norms of reciprocity’, and ‘health and social services’ (α = 0.86). Factor 2 – *Social engagement and communication* comprises five items and is associated with items such as ‘participation in hobby groups’, and ‘participation in volunteer groups’ (α = 0.78). Factor 3 – *Age-friendly physical environment* comprises five items and is strongly associated with items such as ‘barrier-free sidewalks and roads’, ‘barrier-free public facilities’, and ‘parks and trails for exercise and walking’ (α = 0.82). Regarding the correlation between factors, *Social engagement and communication* positively correlated with *Age-friendly physical environment* (r = 0.52, p < 0.001). The factor structure identified by the EFA in the development sample was further validated in the retest sample, exhibiting a similar factorial configuration. The test-retest stability of subscale scores for each factor across the two time points was assessed using Pearson’s correlation coefficient and ICC (Table 3). For the F1 subscale, Pearson’s r was 0.73 (p < 0.001) and the ICC was 0.69 (95% CI: 0.49-0.82). The F2 subscale had Pearson’s r at 0.71 (p < 0.001) and an ICC of 0.67 (95% CI: 0.46-0.81). For the F3 subscale, the coefficients were Pearson’s r = 0.79 (p < 0.001) and an ICC of 0.78 (95% CI: 0.64-0.86).

**Table 1.** Candidate items for age-friendly communities in 2016 and 2019 (23 items, n=61)

| Possible factors within each domain |  | Development samples |  | Retest samples |  |  |
| --- | --- | --- | --- | --- | --- | --- |
|  |  | (2016) |  | (2019) |  |  |
|  |  | Mean (SD) |  | Mean (SD) |  | Difference |
| Parks and sidewalk for exercise and walking | (% many & some) | 73.3 | (10.1) | 73.1 | (11.4) | -0.1 |
| Barrier-free public facilities | (% many & some) | 15.1 | (7.3) | 11.1 | (6.1) | -4.0 |
| Barrier-free trains and buses | (% many & some) | 10.4 | (5.1) | 9.4 | (5.4) | -1.0 |
| Barrier-free sidewalks and road | (% many & some) | 23.0 | (8.1) | 23.2 | (8.6) | 0.3 |
| Stations and bus stops within walking distance | (% many & some) | 24.3 | (10.3) | 21.9 | (11.1) | -2.4 |
| Housing type | (% owned house) | 93.4 | (3.5) | 93.6 | (3.3) | 0.2 |
| Participation in hobby activity group | (% ≥ once a month) | 35.0 | (5.2) | 31.1 | (4.8) | -3.8 |
| Participation in sports groups/clubs | (% ≥ once a month) | 27.6 | (5.1) | 26.7 | (5.0) | -1.0 |
| Participation in learning or cultural group | (% ≥ once a month) | 8.3 | (2.0) | 8.3 | (2.1) | 0.0 |
| Participation in senior citizen club | (% ≥ once a month) | 10.5 | (4.6) | 9.1 | (5.0) | -1.4 |
| Sense of belonging to the community | (% yes & yes, somewhat) | 36.7 | (10.6) | 35.1 | (10.9) | -1.6 |
| Participation in community decisions | (% yes & yes, somewhat) | 42.8 | (11.5) | 35.9 | (11.9) | -6.9 |
| Norms of reciprocity | (% agree strongly & agree) | 53.9 | (6.1) | 55.3 | (6.1) | 1.4 |
| Participation in volunteer group | (% ≥ once a month) | 15.1 | (3.1) | 14.1 | (3.4) | -1.1 |
| Participation in paid work | (% ≥ once a month) | 27.4 | (3.5) | 32.5 | (4.1) | 5.1 |
| Using Internet or email | (% ≥ less than a few imes a month) | 41.1 | (9.1) | 54.2 | (9.6) | 13.0 |
| Frequency of contact with friends | (% ≥ once a month) | 74.3 | (3.7) | 73.5 | (4.9) | -0.8 |
| Health and welfare services | (% yes & yes, somewhat) | 45.0 | (9.4) | 44.0 | (9.1) | -1.0 |
| Awareness of living with dementia | (% yes & yes, somewhat) | 62.8 | (5.3) | 56.5 | (6.7) | -6.3 |
| Participation of people with dementia in<br>community activities | (% yes & yes, somewhat) | 50.7 | (6.9) | 48.1 | (6.5) | -2.6 |
| Understanding of people with dementia | (% yes & yes, somewhat) | 62.1 | (6.0) | 53.4 | (5.7) | -8.7 |
| Decision-making support for people with<br>dementia | (% yes & yes, somewhat) | 10.6 | (3.6) | 13.2 | (4.0) | 2.6 |
| Support for families with dementia | (% yes & yes, somewhat) | 78.5 | (5.8) | 75.5 | (6.5) | -2.9 |

Abbreviations: SD

**Table 2.** Factor loadings of age-friendly communities’ indicators in 2016 and 2019.

|  | Development samples (2016) |  |  | Retest samples (2019) |  |  |
| --- | --- | --- | --- | --- | --- | --- |
|  | Factors 1 | Factors 2 | Factors 3 | Factors 1 | Factors 2 | Factors 3 |
|  | Social<br>inclusion and<br>dementia<br>friendliness | Social<br>engagement<br>and<br>communicatio<br>n | Age-friendly<br>physical<br>environment | Social<br>inclusion<br>and<br>dementia<br>friendliness | Social<br>engagement<br>and<br>communicati<br>on | Age-friendly<br>physical<br>environment |
| Sense of belonging to the community | **0.86** | 0.02 | -0.19 | **0.89** | 0.00 | -0.10 |
| Norms of reciprocity | **0.85** | -0.15 | 0.13 | **0.74** | 0.07 | -0.06 |
| Health and welfare services | **0.79** | 0.11 | 0.03 | **0.58** | -0.03 | 0.25 |
| Participation in community decisions | **0.78** | -0.07 | -0.10 | **0.82** | -0.19 | 0.01 |
| Support for families with dementia | **0.58** | -0.21 | -0.01 | **0.62** | -0.12 | -0.09 |
| Frequency of contact with friends | **0.56** | 0.11 | -0.27 | **0.39** | 0.33 | -0.46 |
| Participation of people with dementia in community activities | **0.56** | 0.26 | 0.21 | **0.58** | 0.10 | 0.12 |
| Participation in hobby activity group | -0.19 | **0.90** | 0.05 | -0.05 | **0.93** | 0.02 |
| Participation in sports groups/clubs | -0.13 | **0.88** | 0.08 | -0.10 | **0.80** | 0.18 |
| Participation in volunteer group | 0.28 | **0.75** | -0.04 | 0.14 | **0.77** | -0.19 |
| Participation in learning or cultural group | 0.20 | **0.63** | 0.09 | 0.05 | **0.64** | 0.00 |
| Using Internet or email | -0.34 | **0.62** | -0.17 | -0.17 | **0.57** | 0.17 |
| Barrier-free sidewalks and roads | -0.14 | -0.28 | **0.99** | 0.31 | -0.06 | **0.91** |
| Barrier-free public facilities | 0.05 | -0.01 | **0.80** | 0.09 | -0.06 | **0.88** |
| Parks and sidewalk for exercise and walking | -0.08 | 0.08 | **0.72** | 0.01 | 0.22 | **0.69** |
| Barrier-free trains and buses | -0.02 | 0.16 | **0.59** | 0.01 | 0.01 | **0.68** |
| Stations and bus stops within walking distance | 0.00 | 0.20 | **0.42** | -0.16 | 0.17 | **0.48** |
| Correlation coefficients between factors |  |  |  |  |  |  |
| Factors 1 | 1.00 | -0.06 | -0.23 | 1.00 | -0.40 | -0.22 |
| Factors 2 |  | 1.00 | 0.52 |  | 1.00 | 0.31 |
| Factors 3 |  |  | 1.00 |  |  | 1.00 |
| $\alpha$ | 0.86 | 0.78 | 0.82 | 0.85 | 0.79 | 0.81 |

**Table 3.** Test-retest reliability.

| | Development samples<br>(2016) | Retest samples<br>(2019) | $r^{\dagger}$ | ICC (95% CI) |
| --- | --- | --- | --- | --- |
|  | Mean (SD) | Mean (SD) |  |  |
| Social inclusion and dementia friendliness | 54.6 (6.2) | 52.9 (6.0) | 0.73* | 0.69 (0.49-0.82) |
| Social engagement and communication | 25.4 (4.0) | 26.9 (4.1) | 0.71* | 0.67 (0.46-0.81) |
| Age-friendly physical environment | 29.2 (6.4) | 27.8 (6.7) | 0.79* | 0.78 (0.64-0.86) |

† Pearson correlation coefficients; \* $P < 0.05$ .
Abbreviations: SD, standard deviation; ICC, intraclass correlation coefficient; CI, confidence interval.

## 4. Discussion

This study developed a 17-item community-level AFC indicator grounded in the WHO AFC guidelines, incorporating considerations of dementia-friendly elements. Furthermore, the validity and reliability of the developed indicator for assessing temporal stability were verified.

This indicator has a three-factor structure. Factor 1 – *Social inclusion and dementia friendliness* covers the WHO AFC core indicators: ‘respect and social inclusion’, ‘communication and information’, and ‘community support and health services’. It also includes principles related to the inclusion of people with dementia. Thus, Factor 1 reflects an inclusive social environment for older people, including those with dementia. Factor 2 – *Social engagement and communication* includes items related to group participation and use of Internet/email. The introduction of Internet and technology use is discussed within the AFC context [28, 29] and the use of Internet suggests a link with enhanced social participation [30]. Thus, Factor 2 assesses the social participation of older people and their use of online communication. This factor covers the following WHO AFC core indicators: ‘social participation’, ‘civic participation and employment’, and ‘communication and information’, where ‘communication and information’ is also included in Factor 1. Factor 3 – *Age-friendly physical environment* covers the WHO core indicators, ‘outdoor spaces and buildings’ and ‘transportation’, and represents the physical environment of older people. Thus, our indicator covers seven of the eight domains of the WHO’s AFC core indicators and includes both the physical and social environment. The Cronbach’s alpha of these factors ranged from 0.78 to 0.86, indicating internal consistency. Furthermore, the reproducibility of this three-factor structure was largely confirmed in a retest sample, albeit with some ambiguity regarding the frequency of meeting friends. Further research with an expanded sample size is needed to validate these findings.

The housing-related items were ultimately excluded from the study. One possible reason for exclusion is that we asked the housing-related questions in terms of housing type and home-ownership status. Given the high rate of owner-occupied housing in this cohort in Japan, this question may not be directly applicable within the AFC framework in the Japanese context. More appropriate housing-related items, such as housing comfort, degree of barrier-free accessibility, and access to community and social services [31, 32], are needed to be identified for inclusion.

Notably, two indicators related to dementia-friendliness remained in the developed indicator, namely ‘support for families with dementia’ and ‘participation of people with dementia in community activities.’ The reason may be their alignment with the domain ‘respect and social inclusion’ in the WHO framework. The excluded dementia-friendly elements, such as ‘awareness of living with dementia’ and ‘understanding people with dementia’ may not necessarily reflect community conditions that include people with dementia, and it may be necessary to measure behaviours and attitudes as well as knowledge items.

The test-retest reliability of the subscale scores was assessed across two time points; the results obtained statistically significant correlation coefficients and ICCs that indicated adequate test-retest reliability (ICC ≥ 0.70) [33]. This suggests that this index is reliable. Therefore, this indicator may give consistent results even when measured at different times.

The strengths of this study are that the indicator focuses on the community-level, incorporates dementia-friendly elements, and has been validated and found reliable in a large number of older people living in various regions of Japan. It is a comprehensive indicator that reflects seven of the eight domains from the WHO framework. However, this study has several limitations. First, because the data is not based on responses from people with physical disabilities, people with dementia, or caregivers, it is unclear whether it completely captures the actual situation in the area. Further research is needed to include such individuals for developing a more comprehensive indicator. Second, this indicator was validated for 61 areas in 16 Japanese municipalities, but does not necessarily have generalizability because it does not cover all of Japan. Expanding the scope of the survey is an important next step to make this indicator applicable to a variety of regions. Finally, while the indicator incorporates dementia-friendly elements, it only partially covers the social environment aspect of DFCs. It also does not fully address the important physical environment features that reduce anxiety and confusion among people with dementia, such as clear colour contrasts in mats, floors; clear directions and street signage; and the placement of street trees and street furniture as navigation aids. [34]. Therefore, this indicator needs further refinement in the future.

## 5. Conclusion

We have developed a community-level age-friendly indicator with sufficient validity and reliability to include the elements of the DFC. It comprises three factors—*Social inclusion and dementia friendliness, Social engagement and communication*, and *Age-friendly physical environment*—aligns with the WHO Framework. The indicator will enable community monitoring, evaluation, and inter-community comparisons, and will help create AFCs and DFCs in rapidly aging Japan.

## Supporting information

etable1

etabel2

## Data Availability

The data underlying this study are from the JAGES and contain sensitive information. Data for research purposes is available upon request. Requests for the JAGES data can be made to.

